# Identifying Barriers to Disclosure to Overcome Barriers to ART Initiation: An explanatory qualitative study on PMTCT in Zambia

**DOI:** 10.1101/2020.11.07.20227637

**Authors:** Tukiya Kanguya, Aybüke Koyuncu, Anjali Sharma, Thankian Kusanathan, Martha Mubanga, Benjamin H. Chi, Michael J. Vinikoor, Mwangelwa Mubiana-Mbewe

## Abstract

**Background:** Though antiretroviral therapy (ART) is widely available, HIV positive pregnant women in Zambia are less likely to start and remain on therapy throughout pregnancy and after delivery. This study sought to understand readiness to start ART among HIV pregnant women from the perspectives of both women and men in order to suggest more holistic programs to support women to continue life-long ART after delivery.

**Methods:** We conducted a qualitative study with HIV positive pregnant women before and after ART initiation, and men with female partners, to understand readiness to start lifelong ART. We conducted a total of 28 in-depth interviews among women and 2 focus group discussions among male partners. Data was transcribed verbatim and analyzed in NVivo 12 using thematic analysis. Emerging themes from the data were organized using the social ecological framework.

**Results:** Men thought of their female partners as young and needing their supervision to initiate and stay on ART. Women agreed that disclosure and partner support were necessary preconditions to ART initiation and adherence and expressed fear of divorce as a prominent barrier to disclosure. Maternal love and desire to look after one’s children instilled a sense of responsibility among women which motivated them to overcome individual, interpersonal and health system level barriers to initiation and adherence. Women preferred adherence strategies that were discrete, the effectiveness of which, depended on women’s intrinsic motivation.

**Conclusion:** The results support current policies in Zambia to encourage male engagement in ART care. To appeal to male partners, messaging on ART should be centered on emphasizing the importance of male involvement to ensure women remain engaged in ART care. Programs aimed at supporting postpartum ART adherence should design messages that appeal to both men’s role in couples’ joint decision-making and women’s maternal love as motivators for adherence.

## Introduction

Despite impressive gains towards reaching global goals for ending HIV, Zambia is short of reaching the 95-95-95 UNAIDS target for HIV diagnosis, antiretroviral therapy (ART) initiation and viral suppression in women. In 2017, approximately 72,000 children where living with HIV and 7,300 children become newly infected with HIV [1]. Data from Southern African region have shown that women who initiated ART during pregnancy had a poor 24-month retention in care, and this contributed to an increasing number of children being infected during the breastfeeding period [2]. Approximately 60% of new HIV infections in children occur during the breastfeeding period due to poor maternal ART adherence and inadequate systems of follow-up for postpartum mothers [3]. Therefore, supporting pregnant women to identify and overcome barriers to long-term ART use is a major priority to eliminate PMTCT in the country.

Barriers to timely ART initiation among HIV pregnant women vary from the fear of the side effects of the drug, lack of spousal involvement to community stigma [2]. Recent research has shown that that the risk factors for low adherence include being of a younger age, less educated and employed in the informal sector [5]. Selected reported barriers for not being ready to start or continue on ART included physical, economic and emotional stresses, depression (especially after delivery), alcohol or drug use, and ART dosing frequency or pill burden [2,4,5]. While many women overcome these barriers during pregnancy, they may disengage from care after birth.

Furthermore, one of the major deterrents to initiation and sustained adherence has been the lack of male involvement in ART care as women need to discuss their HIV status with their husbands before deciding to start ART[6–8]. Men’s attitude to joining Prevention of Mother to Child Transmission (PMTCT) in sub-Saharan Africa has been linked to cultural expectations and the social norm that hold women responsible for attending PMTCT [9]. Men in Botswana regarded antenatal care (ANC) facilities as being “generally unfriendly” to them [10]. Most recently, a review of studies incorporating men suggested that male “support” as well as “involvement” is key to increasing PMTCT uptake [11]. The WHO 2010-2015 guidelines on PMTCT emphasize the need to involve male partners to scale up PMTCT services in sub-Saharan Africa [12]. Despite this, there is limited literature on the perspectives from male partners on reasons women delay to initiate drugs and disengage from care after delivery.

The Zambian government adopted the Elimination of Mother-to-Child Transmission of HIV (EMTCT) goal to eliminate new pediatric HIV infections by 2021 which requires that 95% of HIV positive pregnant women be on treatment and are retained in care [13]. Achieving this goal will require a deeper understanding of the barriers towards both initiation and retention on ART among HIV positive pregnant women from the perspective of both men and women. To address this knowledge gap, we conducted a formative qualitative research study to better understand readiness to start lifelong ART among HIV-infected pregnant women. The research question underlying this study is therefore, Understanding the readiness to start lifelong ART among HIV-infected pregnant women. The findings from this study will be used to design interventions that encourage both male and female involvement in antenatal care in health facilities in Zambia

## Methods

### Research setting

This study was conducted in 2 urban health centers in Lusaka District, Zambia, approximately 1 year after introduction of Option B+ in Zambia (i.e., universal and lifelong ART for all pregnant and breastfeeding women living with HIV). Health centers were purposively sampled in consultation with the Lusaka District Health office to include urban government facilities with medium to high patient volume and physical space for study activities at the clinic. Lusaka district is one of four districts within Lusaka Province, which is home to approximately 20% of Zambia’s population of 17 million [14]. Most residents in Lusaka District live below the poverty line in high-density peri-urban slums or “compounds” with poor access to safe water and sanitation. Similar to many countries in sub-Saharan Africa, the HIV epidemic disproportionately impacts individuals living in urban areas in Zambia with Lusaka province having the highest prevalence of HIV infection among adults (15.7%) [15].

### Study procedures

We utilized formative qualitative research to understand readiness to start lifelong ART among HIV-infected pregnant women. Findings from this research were used to develop and evaluate a quantitative tool to assess ‘readiness’ for ART initiation and an intervention package aimed at supporting adherence and retention on treatment among the pregnant population, for which results are presented elsewhere [16].

This project was reviewed in accordance with CDC human research protection procedures and was determined to be research, but CDC investigators did not interact with human subjects or have access to identifiable data or specimens for research purposes. Ethical approval for the study was obtained from the University of Zambia Biomedical Research Ethics Committee (Lusaka, Zambia) and the University of North Carolina at Chapel Hill Institutional Review Board (Chapel Hill, NC, USA).

### Participants

HIV-infected pregnant women not yet on ART (pre-ART group) and HIV-infected pregnant or postnatal (≤ 42 days after delivery) women on ART (post-ART group) were recruited to participate in in-depth interviews (IDIs). Partners of women who were recently or were currently pregnant were also recruited in the study to participate in focus group discussions (FGDs) in an effort to understand male perspectives on barriers and facilitators of ART initiation and adherence. Male partners may or may not have been the partners of women participating in IDIs, and did not necessarily have partners living with HIV. All participants in the various study groups were 18 years of age or older, did not have a known history of mental illness, were able to communicate in one of the three languages used in the study (English, Nyanja or Bemba) and provided written informed consent to participate in the study. The target sample size needed to reach saturation of themes and patterns emerging from data related to the beliefs, behaviors, and experiences of men and women in each study group was determined prior to sampling and data collection [17,18].

Study participants were recruited using convenience sampling. Women presenting at ANC visits or under-5 clinics at the selected urban health centers were sensitized about the study during the daily routine group health talks. Women interested in the study were screened for study participation. In addition, women were identified through their health records, in collaboration with the health facility staff, and all those meeting inclusion criteria were approached on an individual basis for study participation. All eligible women were recruited after providing informed consent. Sensitization about the study for men was done in the general outpatient clinic, TB clinic or ART clinic at the same health facilities. Men were approached individually to establish interest and those interested were screened for study participation. In addition, community sensitization was done through community leaders; men were then approached individually in the community in various places including markets, bus-stops, churches and other public spaces. Those interested in participating were then invited to the study site and screened for study participation. Eligible men were recruited after providing informed consent. Recruiting of men outside the health facilities was done in order to minimize bias towards men with positive health-seeking behavior.

### Data collection

Data were collected between June and September 2015. Topics shown previously to be important when measuring ART readiness were covered in IDIs, namely: disclosure, partner involvement, psychosocial issues, HIV medication beliefs, and alcohol and drug use [19]. To understand individual barriers and facilitators of ART initiation, we assessed women’s knowledge and understanding of ART, their experiences with stigma and discrimination, their existing support structures, and their individual motivations. HIV-infected pregnant women were enrolled and interviewed before or within seven days of ART initiation (pre-ART group) and then a subset of these women was asked to participate in an additional IDI 2-3 months after their first IDI to assess changes in barriers to initiation over time. Participants who had already initiated ART at presentation (post-ART group) were asked about barriers and facilitators to ART adherence, with topics including: social support, distance from clinics and transportation, cultural norms, partner involvement, patient-provider relationships, HIV-related stigma, and experiences at health facilities. Participants were asked to assess the quality and effectiveness of adherence services already received (e.g. adherence counselling); they were also asked for suggestions for additional or alternative services that could be provided to help improve adherence to ART.

FGDs with men who currently have female partners who are/were recently pregnant concentrated on their knowledge about HIV and PMTCT, factors that impact starting and adhering to ARVs among pregnant women, and the level of men’s engagement in their partner’s healthcare during and after pregnancy defined as support in the home or at clinic visits.

All IDIs and FGDs were conducted in English or one of two local languages (Nyanja and Bemba) and held in a private setting at one of the two study clinics or the Centre for Infectious Disease Research in Zambia (CIDRZ) research facility in Lusaka, Zambia.

### Framework

We used the social ecological framework [20] to organize results by sex. The social ecological model is a theory-based framework for understanding the interactive effects of individual, interpersonal, health system level and structural factors on behavior. The insights provided by the model help design multi-faceted behavior change interventions that went beyond the individual. We defined individual level factors as those within a woman’s control and awareness; interpersonal level factors as a woman’s primary relationships affecting her ART treatment, health system level factors as health care structure and design; and structural level factors as women’s socio-economic environment.

### Analysis

Audio recordings of IDIs and FGDs were transcribed verbatim and translated from local languages of Nyanja and Bemba to English by trained research assistants. Data were analyzed using inductive thematic analysis, which is a comprehensive process involving reading through the transcripts for familiarization and identifying emerging key themes and codes which are then entered into a codebook [21]. The data was subsequently coded by two coders using QSR International’s *NVivo* 12 qualitative data analysis software. Coding was compared amongst the two coders for consistency and similarity. Emerging themes for barriers and facilitators of ART initiation and adherence were organized according to levels of the social ecological framework (individual, interpersonal, health system, or structural level barriers/facilitators).

## Results

### Characteristics of study population

We conducted a total of 28 IDIs among 20 pregnant and postpartum HIV-infected mothers and 2 FGDs among 16 men in groups of 8. Demographic characteristics of the women and men are shown in Tables 1 and 2 respectively. Among 20 women, 12 women had IDIs at only one time-point (5 in pre-ART group, and 7 in post-ART group) and 8 pre-ART women had a follow-up IDI that took place 2-3 months after their first interview. IDIs that occurred only at one time-point were conducted among a mix of pregnant (N=7) and postnatal (≤42 days after delivery) (N=5) women.

**Table 1.**
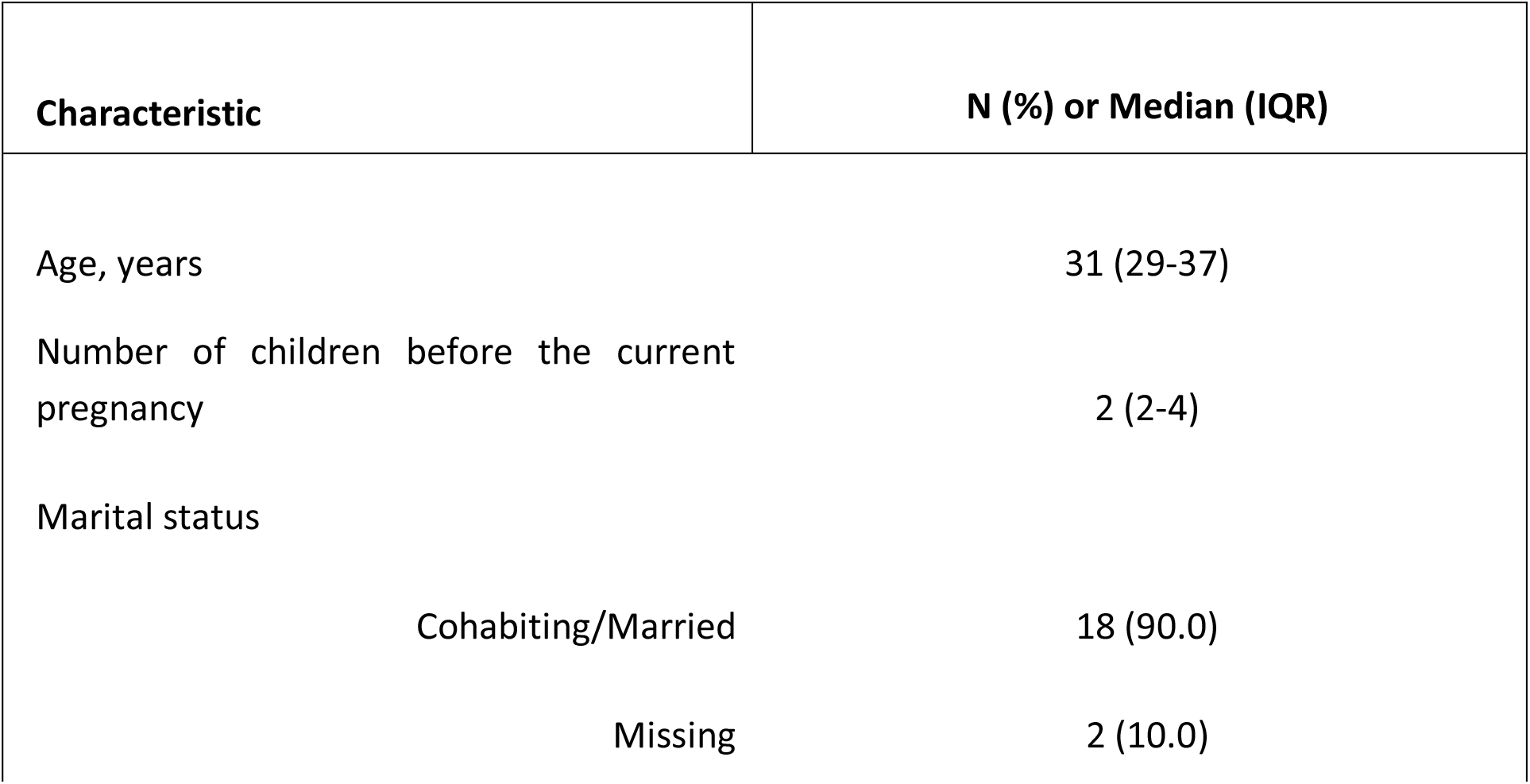
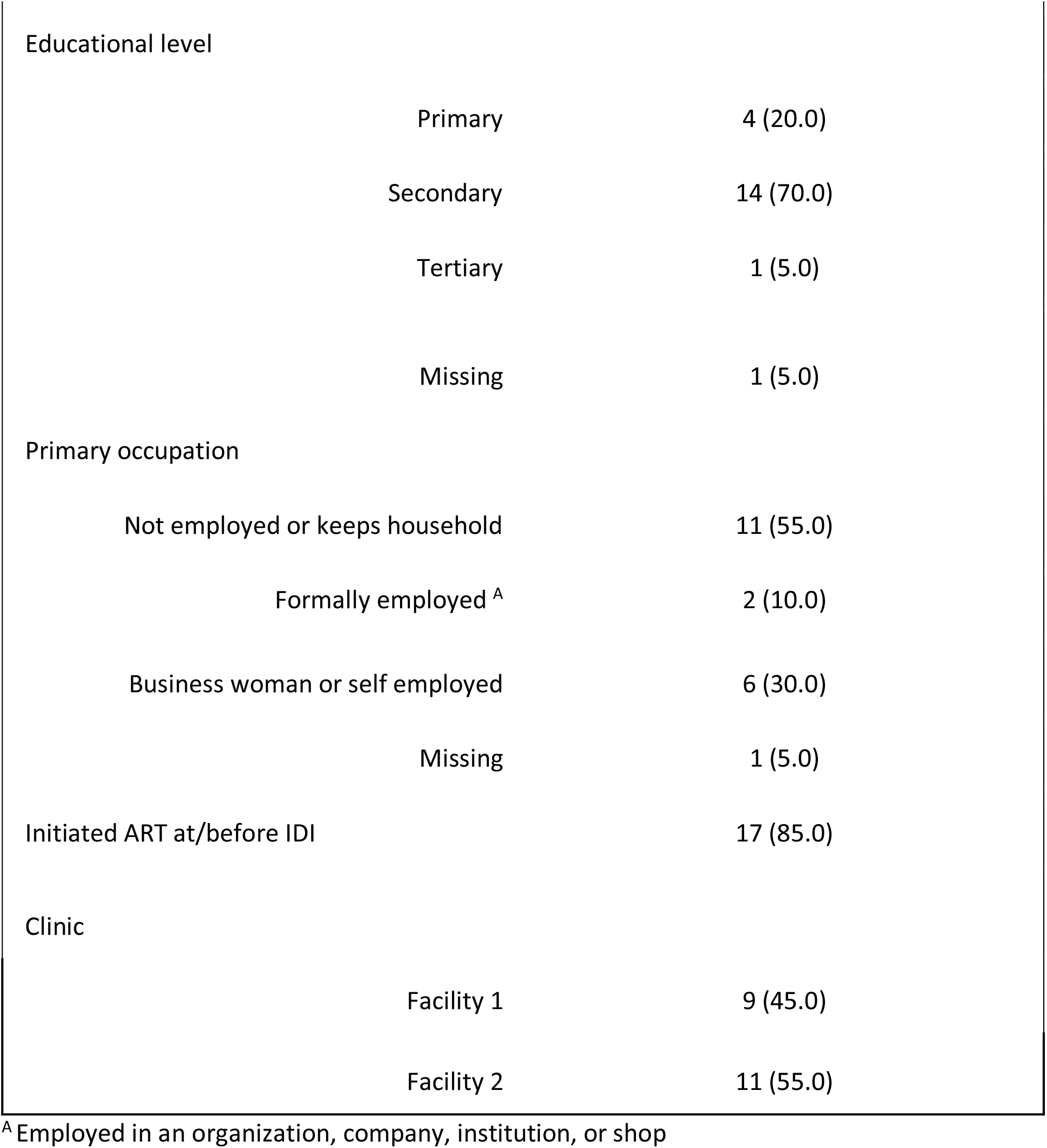
Characteristics of pregnant and postnatal HIV-infected women participating in in-depth interviews, Lusaka, Zambia, 2015.

| Characteristic | N (%) or Median (IQR) |
| --- | --- |
| Age, years | 31 (29-37) |
| Number of children before the current pregnancy | 2 (2-4) |
| Marital status |  |
| Cohabiting/Married | 18 (90.0) |
| Missing | 2 (10.0) |

|  |  |  |
| --- | --- | --- |
| Educational level |  |  |
|  | Primary | 4 (20.0) |
|  | Secondary | 14 (70.0) |
|  | Tertiary | 1 (5.0) |
|  | Missing | 1 (5.0) |
| Primary occupation |  |  |
|  | Not employed or keeps household | 11 (55.0) |
|  | Formally employed <sup>A</sup> | 2 (10.0) |
|  | Business woman or self employed | 6 (30.0) |
|  | Missing | 1 (5.0) |
| Initiated ART at/before IDI |  | 17 (85.0) |
| Clinic |  |  |
|  | Facility 1 | 9 (45.0) |
|  | Facility 2 | 11 (55.0) |

<sup>A</sup> Employed in an organization, company, institution, or shop

**Table 2.**
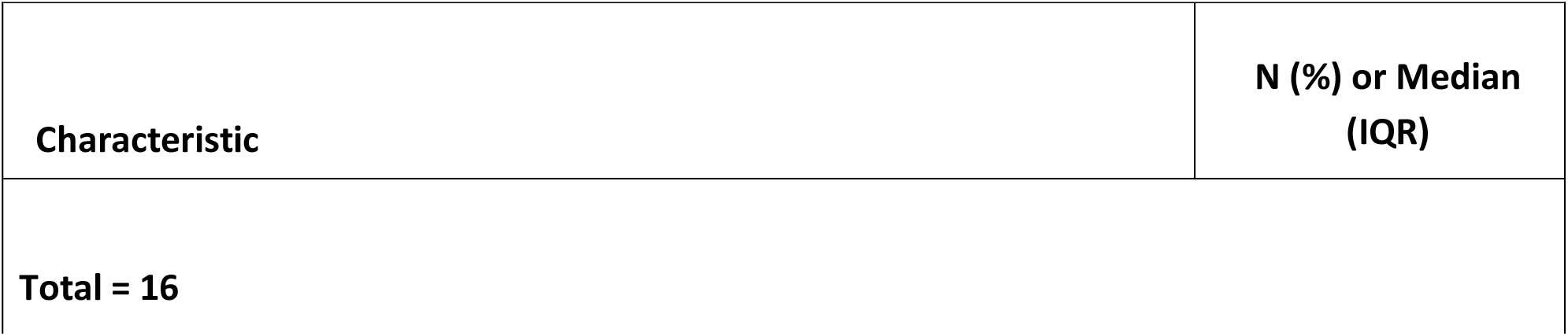
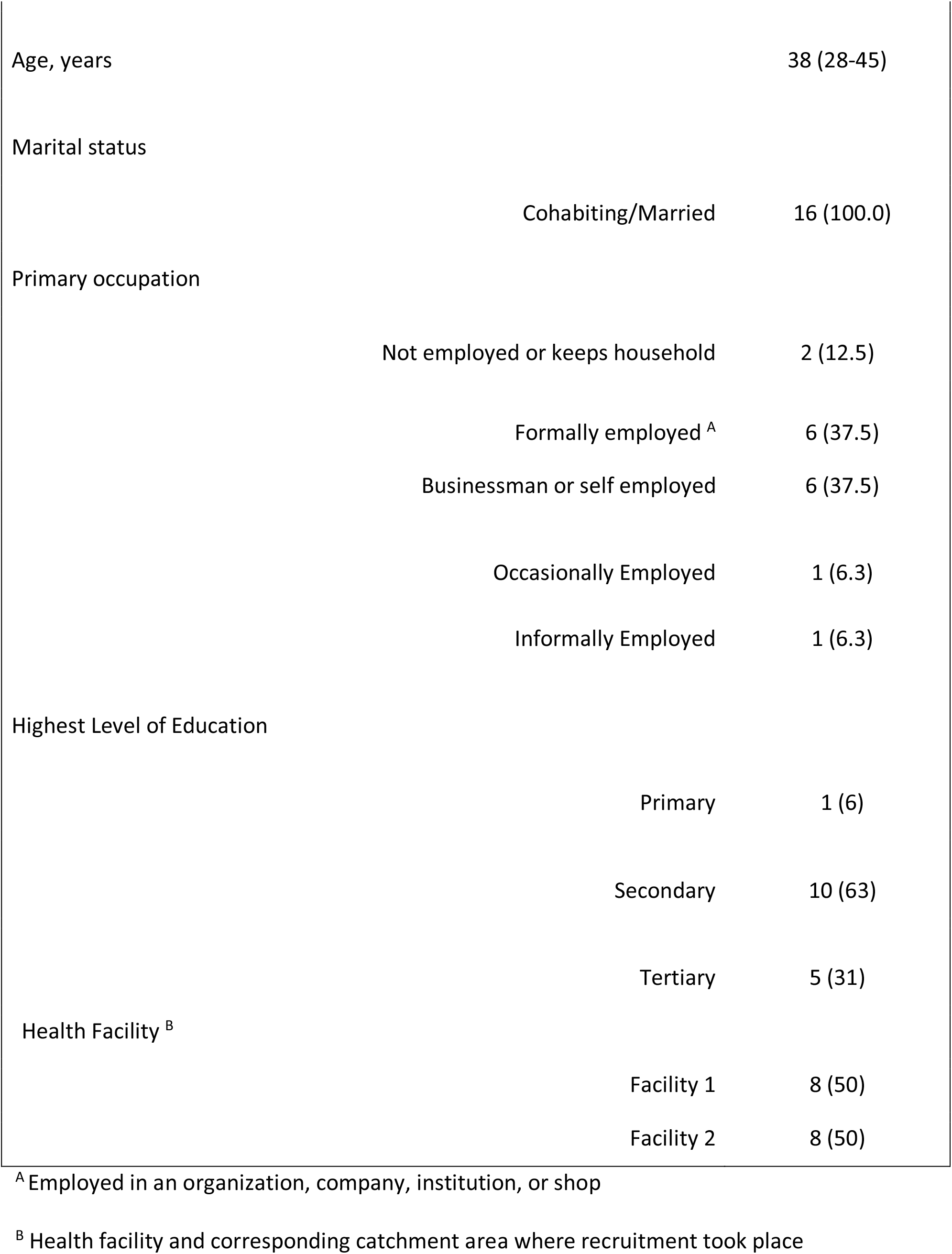
Characteristics of Male partners participating in the Focus Group Discussions (FGDs) in Lusaka, Zambia, 2015.

| Characteristic | N (%) or Median (IQR) |
| --- | --- |
| Total = 16 |  |

|  |  |  |
| --- | --- | --- |
| Age, years |  | 38 (28-45) |
| Marital status |  |  |
|  | Cohabiting/Married | 16 (100.0) |
| Primary occupation |  |  |
|  | Not employed or keeps household | 2 (12.5) |
|  | Formally employed <sup>A</sup> | 6 (37.5) |
|  | Businessman or self employed | 6 (37.5) |
|  | Occasionally Employed | 1 (6.3) |
|  | Informally Employed | 1 (6.3) |
| Highest Level of Education |  |  |
|  | Primary | 1 (6) |
|  | Secondary | 10 (63) |
|  | Tertiary | 5 (31) |
| Health Facility <sup>B</sup> |  |  |
|  | Facility 1 | 8 (50) |
|  | Facility 2 | 8 (50) |

<sup>A</sup> Employed in an organization, company, institution, or shop
<sup>B</sup> Health facility and corresponding catchment area where recruitment took place

Because immediately ART initiation had become standard of care, at the time of their first IDI, 17 of 20 mothers (85.0%) had initiated on ART and 1 mother (4.8%) indicated plans to initiate the day of her interview (Table 1). Only 1 participant who had not initiated ART at the time of their first IDI had a subsequent follow-up IDI, and initiated ART prior to her follow-up interview. All participants with follow-up interviews who had initiated ART at the time of their first IDI (N=7) reported still being on ART at the time of their follow-up interview.

### Women’s perspectives on barriers and facilitators to ART initiation

Pregnant and postnatal women in this study reported a range of barriers to ART initiation within all hierarchies of the social-ecological model (individual, interpersonal, health system, and structural levels). Women who had already initiated ART as well as those who had not yet initiated viewed partner disclosure and partner support of ARVs as necessary for ART initiation. A participant who had not initiated ART at the time of her interview explained:

> *“I always think of how am I going to manage taking ARVs without my partner knowing, because definitely he has to know about me being on treatment. Since I have never disclosed my status, I feel it is unfair for me to start taking ARVs without my partner knowing*.*” (Female, 31 years, had not initiated ART)*.

Barriers to partner disclosure, such as fear of partner blame and fear of divorce, were reported as substantive barriers to ART initiation among both pregnant and postnatal women:

> *“*… *It’s not like I tested HIV positive today, it has been two years. [I have] not disclosed my status to him because there are a lot of fear. He might end up leaving me* … *I think the main challenge that women have is actually disclosure. If I go to a clinic and I am tested alone and am HIV positive, what if he asks me what made me do it? Maybe he would say that, ‘You know what you have been doing, that is why you went for a test’; or he may end up accusing me having extramarital affairs, that is why I went quiet* … *It is very difficult if am tested alone for me to go and tell my partner that I tested positive*.*” (Female, 31 years, had not initiated ART)*.

Only 2 participants indicated that they did not have plans to initiate ART at the time of their interview. Both of these women had not disclosed their status to their partner, and suggested couples testing and counselling as strategies to support partner disclosure. Suggestions for the need for couples testing and counselling were further strengthened by descriptions from delayed ART acceptors (those who did not accept ART the day that they tested HIV positive) for the need to go home and discuss with their partner prior to initiating ART:

> *“You know when you have just been told news like you are HIV positive, it is always hard to accept. So, I had to sit and discuss with my partner; so time passed before we made a decision to start taking ARVs. We finally accepted our status, my partner and I then agreed that I start taking ARVs yesterday*.*” (Female, 32 years, on ART)*.

In the presence of partner disclosure and support of ARVs, wanting to protect the health of the baby and love for one’s children were facilitators to acceptance of one’s HIV status and readiness to initiate ART. While women described wanting to be healthy and prolong one’s own life as other important individual-level motivators for ART initiation, for many participants these individual desires had substantial interactive effects with maternal love. A mother’s love for her unborn child was sufficient to overcome not only individual barriers such as lack of acceptance of one’s status, feeling healthy, lack of confidence in ability to follow ARV regimens, and fear of harm/side-effects due to ARVs, but also interpersonal and community level barriers such as fear of stigma and health system barriers such as long queues at clinics. Maternal desires to protect their children uniquely instilled a self-responsibility to overcome other existing barriers to ART initiation:

> *“You find people who say bad things about HIV positive people who take ARVs. So, it is up to you to ignore all those bad things people say and continue taking ARVs to help yourself. If you take your ARVs you will live long enough to keep your children, do not listen to what people say*.*” (Female, 29 years, on ART)*.

### Women’s perspectives on barriers and facilitators to ART adherence

All women demonstrated proficiency and understanding regarding how ARVs function within the body and the importance of taking doses at the right time. Most women who had initiated ART at the time of the interview expressed no difficulties taking ARV regimens as prescribed by health care workers and only reported few instances of missing doses. Self-reported good adherence, however, was not attributable to a lack of barriers to ART adherence by the women but rather a self-responsibility to overcome these barriers.

When asked about personally experienced or reported barriers to taking ARVs, participants described barriers in all hierarchies of the social-ecological framework. For example, reported individual level barriers included lack of partner disclosure, feeling healthy that belied the need for ARVs, interpersonal and community barriers included difficulties getting permission from work to pick up ARVs and misinformation received from churches, and health system barriers included long queues at the clinic. Despite these barriers, an emergent theme was women’s intrinsic motivation to “be serious” with their ARVs and overcome barriers to adherence. For example, when asked why long queues at a clinic could be a barrier to adherence, one participant explained:

> *“That person may not just be serious about their life because mostly when people who are not serious come to the clinic for appointments and find that there are a lot of people in the queue they will go back. Mostly such people are drunkards and not just serious about their lives, that is what causes them to stop. But if you really want to have a good and healthy life, you can still wait and get your medication*.*” (Female, 23 years, on ART)*

Similar to ART initiation, partner support, and the desire to continue taking care of one’s children and watching them grow were seen as significant motivators for adherence to ART. Strategies suggested by participants to support adherence included partner and family reminders, putting ARVs somewhere in the home you can always see them, and always carrying drugs on one’s person. When asked which one strategy would be best for supporting adherence, even after interviewer suggestions for methods such as follow up by peer educators, SMS reminders, adherence counselling, and material incentives, setting alarms on one’s phone or watch was said to be the best strategy for ART adherence. Alarms were described by participants as an effective strategy for supporting adherence that did not jeopardize confidentiality of one’s HIV status and could thus be utilized both by individuals who had and had not disclosed their HIV status to their partners and peers:

> *“The alarm is the most important thing and if you are scared that people will ask when it rings, you can put it on vibration. The phone is the most effective way for remembering to take ARVs*.*” (Female, 23 years, on ART)*.

Women had diverse opinions on adherence support strategies listed by the interviewer. Some women believed the strategies could be helpful and others believed the strategies would be ineffective or could unintentionally disclose one’s status to partners or community members (SMS reminders and follow-up by peer educators). The opinion that strategies would be ineffective were based on the belief that adherence had to be intrinsically motivated as explained by one participant:

> *“It [incentives] may look as if someone is begging you to live when this is about your life and you just need to encourage yourself*.*” (Female, unknown age, had not initiated ART)*.

### Male partner perspectives on barriers and facilitators to ART initiation among women

Men were very knowledgeable and had a common understanding about HIV care and PMTCT in pregnant women. This knowledge ranged from understanding the need for antenatal care visits, recommended diet while on ART and type of medication that women were supposed to take.

Similar to the women, men reported a range of barriers to ART initiation among the women within hierarchies of the social-ecological model (individual, interpersonal, health system, and structural levels). They highlighted individual barriers to ART initiation such as fear of ARV side effects, self-stigma and spiritual beliefs. However, both men and women agreed that women found it most difficult to overcome individual (e.g., denial) and interpersonal barriers (e.g. fear of divorce or HIV-serodiscordance) to ART initiation and continuation.

### Fear of divorce

Similar to the women’s perspectives, men thought that fear of divorce or spousal separation impeded ART initiation among married women. Both women and men highly valued and guarded their marital relationship. Having a healthy and positive relationship with their spouse superseded individual physical health. Men repeatedly reported that women were afraid of the marital repercussions of telling them their status and as a result would not take ARVs:

> *“They fear being divorced and also fear the rejection by families*.*” (Male, 39 years)*
>
> *“Another thing is the fear of being divorced. They may say that if I have not disclosed to my husband and start taking ARVs without him knowing, the marriage will come to the end. So mostly they want to protect their marriage, they do not want to be divorced and this is why they do not feel ready to start taking their drugs*.*” (Male, 29 years)*

### Discordant relationships

Most of the HIV negative men in discordant relationships reported that their female partners suffered emotional distress. Women in discordant relationships viewed themselves as being marginalized and struggled with understanding how they acquired HIV despite being faithful; these feelings delayed their acceptance of ART. One man thought that women may feel more emotionally supported if their partner was HIV positive:

> *“On the other hand, I still feel psychologically that she would have preferred it if both of us were HIV positive and taking drugs because she feels marginalized. [Our sero-discordant status] is something that even I have failed to understand, it’s just God’s doing. I have no words to ask why and why not because God himself knows better*.*” (Male, 22 years)*

### Preference for antenatal HIV testing

In Zambia, the HIV testing available options are routine provider-initiated testing (STI, antenatal and TB clinics), Voluntary Counselling and Testing, Community Based (door to door, mobile outreach) and diagnostic testing. Many men reported that women preferred testing for HIV in antenatal clinics rather than in ART clinics because they thought of them as more private and comfortable. Majority of the men reported that their partners tested for HIV at antenatal visits, preventing them from HIV testing as a couple. Because the men were initially not aware of their partner’s status it was difficult for them to be involved in the health care of partner and also to provide support.

> *“They don’t want to have their HIV test done at voluntary counselling and testing (VCT) so that they know if they are positive or negative but they prefer it done at antenatal. It’s like that’s where they feel comfortable*.*” (Male, 39 years)*
>
> *“Women don’t want to go for VCT so the best thing to do is to accompany them*.*’’ (Male, 29 years)*

### Male partner perspectives on barriers and facilitators to ART adherence among women

Similar to initiation, men reported facilitators to adherence at both individual and interpersonal level of the social ecological framework. Men reported that strengthening facilitators at individual and interpersonal level improved adherence among the women. At the individual level, men suggested alarm reminders and at the interpersonal level, partner supervision and support from immediate family as facilitators to adherence.

### Alarm reminders

Similar to the findings from the women, men perceived alarm reminders as helpful in improving adherence among pregnant women. The men reported that they would set alarms on their phones to remind their partners to take medication. Having a fixed time set for taking medication as a couple improved ART uptake among the women:

> *“On the other hand you have to put a reminder on the phone; an alarm which when it rings, reminds you to take your drugs*.*” (Male, 21 years)*

### Paternalistic supervision

According to the men, women needed to be guided by them and take ART under supervision, saying that, “men were the lions and women the sheep”. They reported that men led women to start and remain on ARVs because women went through a phase of constant denial of their HIV diagnosis.

> *“Women also behaves like children. If you give them medicine, some hide them under the pillow or even throw them away and sometimes they complain that from the time they started taking the same medicine they have never seen any change. So monitoring is required*.*” (Male, 32 years)*

### Immediate family support

At the interpersonal level, the involvement of the family/household in the uptake of medication helped improve adherence to ART by HIV positive women. Members of the family that were knowledgeable/aware of the status of the women would physically remind them to take their medication. Men believed they were the first source of support in helping to improve adherence, followed by children in the household.

> *“The strategies -- firstly it is disclosure, upon disclosing to your husband or children, they need know what time you will be taking the drugs so that they can remind you in case you forget” (Male, 21 years)*
>
> *“There are also psychological reminders like if you have a daughter, you may not disclose to her the status of her mother but just tell her that your mother will be taking BP medication and you should be reminding her every day. You even tell her that this is the responsibility that I have given you and maybe even give her a certain amount of money for lunch at school. That is just a psychological way but wherever she is she will be reminded that she has to remind her mother to take her medication*.*” (Male, 22 years)*

## Discussion

Our study identified partner disclosure as a critical facilitator for maternal ART initiation among Lusaka participants. The presence of partner disclosure and support at the interpersonal level was seen as sufficient to overcome all other self-reported socio-ecological barriers to ART initiation. While both women and men also identified male partners as facilitators of ART adherence, they valued different types of support. Men emphasized constant supervision and leadership to ensure women adhere to ART. Women, on the other hand, emphasized the need for open and non-punitive interactions. These results add to research in other low and middle-income countries (LMIC) offering a more nuanced understanding of partner support to help guide appropriate messaging to increase uptake of Option B+.

Lack of male knowledge on HIV and PMTCT has previously been identified in numerous studies in sub-Saharan Africa as a barrier to male involvement in PMTCT, partner disclosure, and women’s ART initiation and adherence [22–26]. The high knowledge levels on HIV, PMTCT, and benefits of ARVs among men in our study simultaneously underscore reach of PMTCT promotion but also the inadequacy of the messaging in tailoring male involvement to the expressed needs of their HIV positive pregnant and breastfeeding partners. Our results suggest that interventions aimed at addressing barriers to disclosure, initiation, and adherence could benefit from appealing to the value both men and women place on their relationship as a strategy to increase male involvement in joint decision-making for women’s HIV care. Our findings utilize data from 2015, therefore barriers and facilitators for ART initiation and adherence among pregnant and postpartum women may have subsequently changed. Reaching the last 5-10% of women in PMTCT elimination goals will require creative strategies. However, interventions targeting the barriers identified in this analysis have not been widespread. Innovative methods focused on the recruitment of men as pioneers in women’s ART initiation and adherence may still be a relevant and necessary strategy for addressing fear of divorce and partner blame as barriers among women who have not yet initiated ART, decreasing delayed acceptance of ART, and supporting ART adherence among pregnant and postpartum women.

Our results complement existing literature identifying partner disclosure and support as consistently important facilitators of ART initiation among pregnant women [22–30], but are the first to suggest disclosure/support are both necessary and sufficient to overcome all other self-reported barriers to initiation when coupled with a mother’s love for her children. While participants in our study acknowledged similar barriers to ART initiation identified by existing research in sub-Saharan Africa such as community stigma and feeling healthy, in the presence of partner support, participants reported both self-efficacy and responsibility to overcome these barriers mediated by their love for their children. The narrative of self-efficacy in overcoming barriers to ART initiation and adherence was also reflected in the selection of phone-based alarms by both women and men in our study as an effective method for supporting ART adherence.

Contrary to previous research conducted among people living with HIV (PLHIV) in Zambia in 2010-11, dissatisfaction with HIV counselling and misinformation about PMTCT/ARVs were not prominent barriers to ART initiation among women in our study population [31]. These findings may be suggestive of improvements to community sensitization and HIV counselling efforts in Zambia. The scale up of HIV care services and ARV delivery in Zambia is further reflected by contrasting the lack of prominent structural level barriers to ART initiation by pregnant mothers in our study to the views of PLHIV in Lusaka in 2010 who reported fear of long-term availability of treatment as a significant barrier to ART initiation [31].

Our finding that love for one’s children and desire to have healthy HIV-free babies were the main motivators for ART initiation and adherence is consistent with existing research [22,24,25,28,30]. However, existing concerns that maternal love may have less influence on ART adherence if the baby is HIV-negative at birth and/or after the breastfeeding period were not supported in our study [23].. Our findings suggest that women may continue on ART in order to live long healthy lives and to take care of their children and watch them grow.

Our ability to draw conclusions regarding the strength of a mother’s love for her children as a long-term facilitator of ART adherence are limited given that women enrolled in our study were either currently pregnant or ≤ 42 days postpartum at the time of their IDI; however, most of the women already had other children at the time of the interview. Our findings may also have limited generalizability given that women could only be recruited for our study if already attending ANC/under-5 clinics and exhibiting positive health seeking behavior related to their child’s health. Our study’s reliance on self-reported adherence may also limit our ability to identify barriers to ART adherence due to social desirability bias and women’s unwillingness to admit poor adherence, which may have contributed to the low prevalence of self-reported poor adherence in our study. Findings from similar qualitative research among HIV-infected pregnant women in sub-Saharan Africa have identified heterogeneity in self-reported adherence, suggesting that challenges with ART adherence may still be identified in the context of self-report [28]. Despite this, prospective follow-up with HIV-infected mothers after the breastfeeding period with more rigorous measurement of ART adherence is needed to confirm the findings of this study. Additional research is also needed to confirm the findings of this analysis given that our results may have limited generalizability to rural areas though increasing migration of rural populations into urban cities in the last decade may increase the relevance of our findings to rural areas in Lusaka and sub-Saharan Africa [32,33].

## Conclusion

While women reported barriers to ART initiation and adherence at all four levels of the social-ecological framework, interventions at the health systems level and structural level may have limited capabilities to facilitate ART initiation and adherence if they do not also address interpersonal barriers related to partner disclosure and support. In order to realize Zambia’s ambitious pediatric HIV elimination goals, PMTCT programs need to strongly promote male engagement and utilize behavioral interventions that address the real and perceived negative consequences of HIV status disclosure. Designing messaging that appeal to men’s role in couples’ joint decision-making for household healthcare and to women’s maternal love may motivate women to continue on ART after delivery.

## Data Availability

Data is accessible on request to: Chief Scientific Officer, P.O. Box 34681, Centre for Infectious Disease Research (CIDRZ), Lusaka, Zambia

## Acknowledgements

The authors thank the study participants and the Centre for Infectious Disease Research in Zambia (CIDRZ) staff for their important contributions.

## Notes

### Competing Interest Statement

The authors have declared no competing interest.

### Funding Statement

This research has been supported by the President's Emergency Plan for AIDS Relief through the United States Centers for Disease Control and Prevention (CDC) under the terms of cooperative agreement number U01GH000530 (Funding recipient MMM). The funders had no role in study design, data collection and analysis, decision to publish, or preparation of the manuscript. The findings and conclusions in this report are those of the authors and do not necessarily represent the official position of the funding agencies.

### Author Declarations

Ethical approval for the study was obtained from the University of Zambia Biomedical Research Ethics Committee (Lusaka, Zambia) and the University of North Carolina at Chapel Hill Institutional Review Board (Chapel Hill, NC, USA).

## References

1. Joint United Nations Programme on HIV/AIDS (UNAIDS), UNAIDS D. Programme on HIV. AIDS. 2018; 2018:376p. Available from: https://www.unaids.org/sites/default/files/media_asset/unaids-data-2018_en.pdf

2. Knettel BA, Cichowitz C, Ngocho JS, Knippler ET, Chumba LN, Mmbaga BT, Watt MH. Retention in HIV care during pregnancy and the postpartum period in the Option B+ era: a systematic review and meta-analysis of studies in Africa. Journal of acquired immune deficiency syndromes (1999). 2018 Apr 15;77(5):427. doi:10.1097/QAI.0000000000001616

3. UNAIDS. 2015 Progress Report on the Global Plan towards the elimination of new HIV infections among children and keeping their mothers alive. 2015. Available: https://www.unaids.org/sites/default/files/media_asset/JC2774_2015ProgressReport_GlobalPlan_en.pdf

4. Minja L, Cichowitz C, Knettel BA, Mahande MJ, Kisigo G, Knippler ET, Ngocho JS, Mmbaga BT, Watt MH. Attitudes Toward Long-Term Use of Antiretroviral Therapy Among HIV-Infected Pregnant Women in Moshi, Tanzania: A Longitudinal Study. AIDS and Behavior. 2019 Sep 1;23(9):2610-7. doi.org/10.1007/s10461-019-02622-5

5. Kiwanuka G, Kiwanuka N, Muneza F, Nabirye J, Oporia F, Odikro MA, et al. Retention of HIV infected pregnant and breastfeeding women on option B+ in Gomba District, Uganda: a retrospective cohort study. BMC Infectious Diseases. 2018;18: 533. doi:10.1186/s12879-018-3450-9

6. Hershow RB, Zimba CC, Mweemba O, Chibwe KF, Phanga T, Dunda W, Matenga T, Mutale W, Chi BH, Rosenberg NE, Maman S. Perspectives on HIV partner notification, partner HIV self-testing and partner home-based HIV testing by pregnant and postpartum women in antenatal settings: a qualitative analysis in Malawi and Zambia. Journal of the International AIDS Society. 2019 Jul;22:e25293. doi.org/10.1002/jia2.25293

7. Orne-Gliemann J, Tchendjou PT, Miric M, Gadgil M, Butsashvili M, Eboko F, Perez-Then E, Darak S, Kulkarni S, Kamkamidze G, Balestre E. Couple-oriented prenatal HIV counseling for HIV primary prevention: an acceptability study. BMC public health. 2010 Dec 1;10(1):197. doi.org/10.1186/1471-2458-10-197

8. Theuring S, Mbezi P, Luvanda H, Jordan-Harder B, Kunz A, Harms G. Male involvement in PMTCT services in Mbeya Region, Tanzania. AIDS Behav. 2009;13 Suppl 1: 92–102. doi:10.1007/s10461-009-9543-0

9. Morfaw F, Mbuagbaw L, Thabane L, Rodrigues C, Wunderlich A-P, Nana P, et al. Male involvement in prevention programs of mother to child transmission of HIV: a systematic review to identify barriers and facilitators. Syst Rev. 2013;2: 5. doi:10.1186/2046-4053-2-5

10. Auvinen J, Suominen T, Välimäki M. Male participation and prevention of human immunodeficiency virus (HIV) mother-to-child transmission in Africa. Psychol Health Med. 2010;15: 288–313. doi:10.1080/13548501003615290

11. Takah NF, Atem JA, Aminde LN, Malisheni M, Murewenhema G. The impact of approaches in improving male partner involvement in the prevention of mother-to-child transmission of HIV on the uptake of safe infant feeding practices by HIV positive women in sub-Saharan Africa. A systematic review and meta-analysis. PloS one. 2018 Dec 3;13(12):e0207060. doi.org/10.1371/journal.pone.0207060

12. Millennium Development Goals, United Nations, General Assembly, Special Session, World Health Organization. PMTCT strategic vision 2010-2015: preventing mother-to-child transmission of HIV to reach the UNGASS and Millennium Development Goals?: moving towards the elimination of paediatric HIV. Geneva: World health organization; 2010. Available: http://www.who.int/hiv/pub/mtct/strategic_vision.pdf

13. The Republic of Zambia Ministry of Health. Elimination of Mother-to-Child Transmission of HIV and Syphilis National Operational Plan 2019-2021. Available: https://www.moh.gov.zm/wp-content/uploads/filebase/Elimination-of-Mother-To-Child-Transmission-of-HIV-and-Syphilis-National-Operation-Plan-2019-2021.pdf

14. Lusaka (Province, Zambia) - Population Statistics, Charts, Map and Location. [cited 14 Oct 2019]. Available: https://www.citypopulation.de/php/zambia-admin.php?adm1id=05

15. Ministry of Health, Zambia. Zambia Population-based HIV Impact Assessment (ZAMPHIA) 2016: Final Report. Lusaka, Ministry of Health. 2019 Feb.

16. Mubiana-Mbewe M, Bosomprah S, Kadota JL, Koyuncu A, Kusanathan T, Mweebo K, et al. Effect of Enhanced Adherence Package on Early Retention in Care Among HIV-Positive Pregnant Women in Zambia: An Individual Randomized Controlled Trial. Rochester, NY: Social Science Research Network; 2019 Oct. Report No.: ID 3467001. Available: https://papers.ssrn.com/abstract=3467001

17. LeCompte MD, Schensul JJ. Designing & conducting ethnographic research. Walnut Creek, Calif: AltaMira Press; 1999.

18. Krueger RA, Casey MA. Focus groups: a practical guide for applied research. 4th ed. Los Angeles: SAGE; 2009.

19. Development of an Easy to Use Tool to Assess HIV Treatment Readiness in Adolescent Clinical Care Settings. [cited 23 Nov 2019]. Available: https://www.ncbi.nlm.nih.gov/pmc/articles/PMC3203751/

20. Golden SD, Earp JAL. Social ecological approaches to individuals and their contexts: twenty years of health education & behavior health promotion interventions. Health Educ Behav. 2012;39: 364–372. doi:10.1177/1090198111418634

21. Nowell LS, Norris JM, White DE, Moules NJ. Thematic Analysis: Striving to Meet the Trustworthiness Criteria. International Journal of Qualitative Methods. 2017;16: 1609406917733847. doi:10.1177/1609406917733847

22. Hodgson I, Plummer ML, Konopka SN, Colvin CJ, Jonas E, Albertini J, et al. A Systematic Review of Individual and Contextual Factors Affecting ART Initiation, Adherence, and Retention for HIV-Infected Pregnant and Postpartum Women. PLoS One. 2014;9. doi:10.1371/journal.pone.0111421

23. Chadambuka A, Katirayi L, Muchedzi A, Tumbare E, Musarandega R, Mahomva AI, et al. Acceptability of lifelong treatment among HIV-positive pregnant and breastfeeding women (Option B+) in selected health facilities in Zimbabwe: a qualitative study. BMC Public Health. 2017;18. doi:10.1186/s12889-017-4611-2

24. Kim MH, Zhou A, Mazenga A, Ahmed S, Markham C, Zomba G, et al. Why Did I Stop? Barriers and Facilitators to Uptake and Adherence to ART in Option B+ HIV Care in Lilongwe, Malawi. PLoS One. 2016;11. doi:10.1371/journal.pone.0149527

25. Elwell K. Facilitators and barriers to treatment adherence within PMTCT programs in Malawi. AIDS Care. 2016;28: 971–975. doi:10.1080/09540121.2016.1153586

26. Naigino R, Makumbi F, Mukose A, Buregyeya E, Arinaitwe J, Musinguzi J, Wanyenze RK. HIV status disclosure and associated outcomes among pregnant women enrolled in antiretroviral therapy in Uganda: a mixed methods study. Reproductive Health. 2017 Dec 1;14(1):107. doi.org/10.1186/s12978-017-0367-5

27. Katirayi L, Namadingo H, Phiri M, Bobrow EA, Ahimbisibwe A, Berhan AY, et al. HIV-positive pregnant and postpartum women’s perspectives about Option B+ in Malawi: a qualitative study. J Int AIDS Soc. 2016;19. doi:10.7448/IAS.19.1.20919

28. Buregyeya E, Naigino R, Mukose A, Makumbi F, Esiru G, Arinaitwe J, et al. Facilitators and barriers to uptake and adherence to lifelong antiretroviral therapy among HIV infected pregnant women in Uganda: a qualitative study. BMC Pregnancy Childbirth. 2017;17. doi:10.1186/s12884-017-1276-x

29. Gill MM, Umutoni A, Hoffman HJ, Ndatimana D, Ndayisaba GF, Kibitenga S, et al. Understanding Antiretroviral Treatment Adherence Among HIV-Positive Women at Four Postpartum Time Intervals: Qualitative Results from the Kabeho Study in Rwanda. AIDS Patient Care and STDs. 2017;31: 153–166. doi:10.1089/apc.2016.0234

30. Patel RC, Odoyo J, Anand K, Stanford-Moore G, Wakhungu I, Bukusi EA, et al. Facilitators and Barriers of Antiretroviral Therapy Initiation among HIV Discordant Couples in Kenya: Qualitative Insights from a Pre-Exposure Prophylaxis Implementation Study. PLOS ONE. 2016;11: e0168057. doi:10.1371/journal.pone.0168057

31. Musheke M, Bond V, Merten S. Deterrents to HIV-patient initiation of antiretroviral therapy in urban Lusaka, Zambia: a qualitative study. AIDS Patient Care STDS. 2013;27: 231–241. doi:10.1089/apc.2012.0341

32. Hommann K, Lall SV. Which Way to Livable and Productive Cities?: A Road Map for Sub-Saharan Africa. The World Bank; 2019. doi:10.1596/978-1-4648-1405-1

33. The World Bank. Republic of Zambia Systematic Country Diagnostic. 2018 Mar. Report No.: 124032-ZM.

